# Virtual Pooling Enables Accurate, End-to-End Multi-Institutional Study Execution and Causal Inference Without Centralized Data Sharing

**DOI:** 10.64898/2026.03.24.26349123

**Authors:** Ishtiyaque Ahmad, Aryan Ayati, Kunlong Liu, Stella Ko, Nicole Bonine, David Tabano, Nina Malik, Tianchu Lyu, Kai Zheng, Vivek A. Rudrapatna, Trinabh Gupta

## Abstract

**Background:** Multicenter retrospective studies often rely on bringing patient-level data together into a single repository, introducing substantial regulatory and operational barriers. Federated analytics provides a privacy-preserving alternative; however, existing implementations are complex to use, require extensive manual effort for data cleaning, preprocessing, and harmonization, and produce approximate rather than ground-truth results for many biostatistical methods. *Virtual Pooling* (VP) is a recently developed multicenter study execution platform designed to overcome these limitations. In this study, we evaluate whether VP can replicate a published multicenter retrospective study end-to-end—including data preprocessing, regression analysis, and causal inference—without centralized data aggregation.

**Methods:** We deployed VP at the University of California, San Francisco (UCSF) and the University of California, Irvine (UCI) and attempted to replicate a published study of diabetic eye disease screening practices (UCSF N = 2,592; UCI N = 5,642). VP supported all phases of this two-center study, including data cleaning, harmonization, feature engineering, imputation, propensity score estimation, patient matching, and model estimation, all conducted through a single interface without manual coordination between centers. We verified preprocessing correctness and compared descriptive statistics and causal effect estimates with those from the original study, which relied on data transfers across the centers. We also measured the latency overhead introduced by VP.

**Results:** VP was deployed without hospital infrastructure changes, new or non-standard governance agreements, or dedicated IT support. All preprocessing steps executed correctly, with individual preprocessing operations and descriptive statistics completing in under 1 second, logistic regression in under 10 seconds, and propensity score matching in under 30 seconds. Descriptive statistics for all 30 baseline covariates were numerically identical to the original study. Univariate regression results identifying predictors of completed screening were also identical, with recent eye clinic referral (OR = 56.7; 95% CI: 42.1–76.4) and history of eye disease (OR = 6.4; 95% CI: 5.6–7.4) as the strongest predictors. VP also reproduced pooled causal estimates of automated referrals, showing an increase in screening completion from 21% to 36% at UCSF and from 13% to 34% at UCI.

**Conclusion:** VP enables accurate, end-to-end multicenter clinical studies without centralized data sharing. By providing a single interface that supports the full analytical workflow, from uncleaned and unharmonized data through statistical results, and by exactly reproducing pooled results, VP eliminates manual coordination and data transfers across centers. These findings validate its practical potential to transform multicenter retrospective studies, particularly in contexts where data sharing is time-consuming, bureaucratic, or restricted.

## Introduction

High-quality and generalizable medical research relies on data drawn from large, diverse populations^1^. Despite the widespread adoption of electronic health records (EHRs), pooling patient data across institutions remains a challenge. Centralized datasets from multiple health centers remain the gold standard to facilitate methodologically rigorous statistical analysis and machine learning, providing comprehensive access to diverse patient data^2^. However, their implementations are often obstructed by legal, ethical, and technical barriers—ranging from patient privacy/confidentiality concerns and institutional data governance restrictions to administrative burdens involving consent, data use agreements, and de-identification procedures. Even when these hurdles are overcome, centralized datasets are costly to maintain and carry residual privacy risks, as any breach exposes the full dataset to potential misuse^3^.

To address these challenges, several privacy-preserving approaches enable collaborative analysis without direct data sharing, including meta-analysis of site-level results, distributed regression^4,5^, and federated analytics^6,7^. However, these approaches typically assume that data are already cleaned and harmonized across sites^8^, whereas in practice multicenter EHR data are often heterogeneous and noisy, requiring extensive manual, site-specific cleaning and coordination. Even after harmonization, many approaches do not support complex, multi-step biostatistical analyses or produce only approximate results^9–12^, particularly when data distributions differ across centers^4,13–16^. Finally, many existing multicenter analysis tools are difficult to use, restrict iterative data exploration by requiring complete analysis pipelines upfront^17–19^, and place a heavy burden on researchers to understand and manage underlying technical infrastructure^9,20–23^.

We recently developed virtual pooling (VP), a lightweight, multicenter study execution platform to address these limitations^24–26^. VP provides a single, user-friendly analytical interface that allows researchers to perform a broad range of statistical and machine learning analyses on siloed datasets while keeping patient-level data securely at each center and abstracting away center-specific infrastructure, deployment, and operational complexity. VP supports all phases of a typical retrospective EHR-based study—including data cleaning, harmonization, feature engineering, imputation, propensity score estimation, patient matching, and model estimation— without manual coordination across centers. Researchers interact with VP through a single, interactive analytical interface that supports incremental code development and real-time inspection of intermediate results, mirroring the experience of analyzing a centralized dataset without requiring a fully predefined workflow. VP also produces results identical to those obtained from centralized analyses and it integrates with existing governance workflows and infrastructure at heterogeneous institutions.

To date VP has been validated in simulated settings and for individual estimation routines such as Cox proportional hazards regression ^24–26^; however, it has not yet been tested in real multicenter deployments involving a full end-to-end pipeline including data preprocessing and harmonization. In this work, we aim to present the first rigorous validation of VP by deploying it across two academic health systems (UCSF and UCI) and comparing its end-to-end workflow and results to a recently published multicenter EHR-based study of diabetic eye disease screening^27^. In doing so, we hope to show that VP is practical, accurate, easy-to-use, and ready to support many multicenter, retrospective studies across diverse healthcare institutions, especially in contexts where data sharing is time-consuming, paperwork-intensive, or impractical.

## Methods

### Study Design and Population

We conducted a retrospective observational study that followed the design and analysis of a previously published study on barriers to diabetic eye disease screening at UCSF and UCI. Like the original study^27^, eligible patients had a diagnosis of type 2 diabetes mellitus (T2DM) and at least one primary care visit between January 1, 2020, and December 31, 2022. This study period was chosen to allow up to two years of follow-up for all patients, with follow-up extending through 2024. Clinical data were extracted from local EHR systems and retained within each institution’s secure environment. We analyzed 30 covariates aligned with the original study, including demographic, clinical, and neighborhood-level social determinants of health. Outcomes included screening referral and completion, diabetic retinopathy diagnosis within one year, and treatment within two years.

### Virtual Pooling (VP) Framework

We used VP, a privacy-preserving multicenter study execution platform designed to support the full end-to-end lifecycle of a typical EHR-based retrospective study while eliminating the needs for transferring patient-level data across institutions. Unlike existing federated and distributed approaches that assume pre-cleaned and harmonized data, VP natively supports early-stage study tasks such as data cleaning, harmonization, feature engineering, and missing data, in addition to downstream statistical and machine learning analyses. Researchers interact with VP through a single and interactive analytical interface, writing analysis code and seeing results as they would for a centralized dataset, while patient-level data remains secure at each center. This design enables analysts to write and refine code incrementally, inspect intermediate results, and adapt analysis plans in real-time, rather than predefine and submit an entire analysis workflow upfront, as required by many existing systems^17–19^. VP also abstracts site-specific infrastructure and coordination, allowing analysts to focus on study design and inference, and is designed to produce results equivalent to centralized analyses while complying with institutional governance constraints.

We deployed VP (v1.1 as of October 2024) as shown in Figure 1. VP consists of two tightly integrated components that work together to enable secure, federated data analysis across institutions:

**Figure 1:**
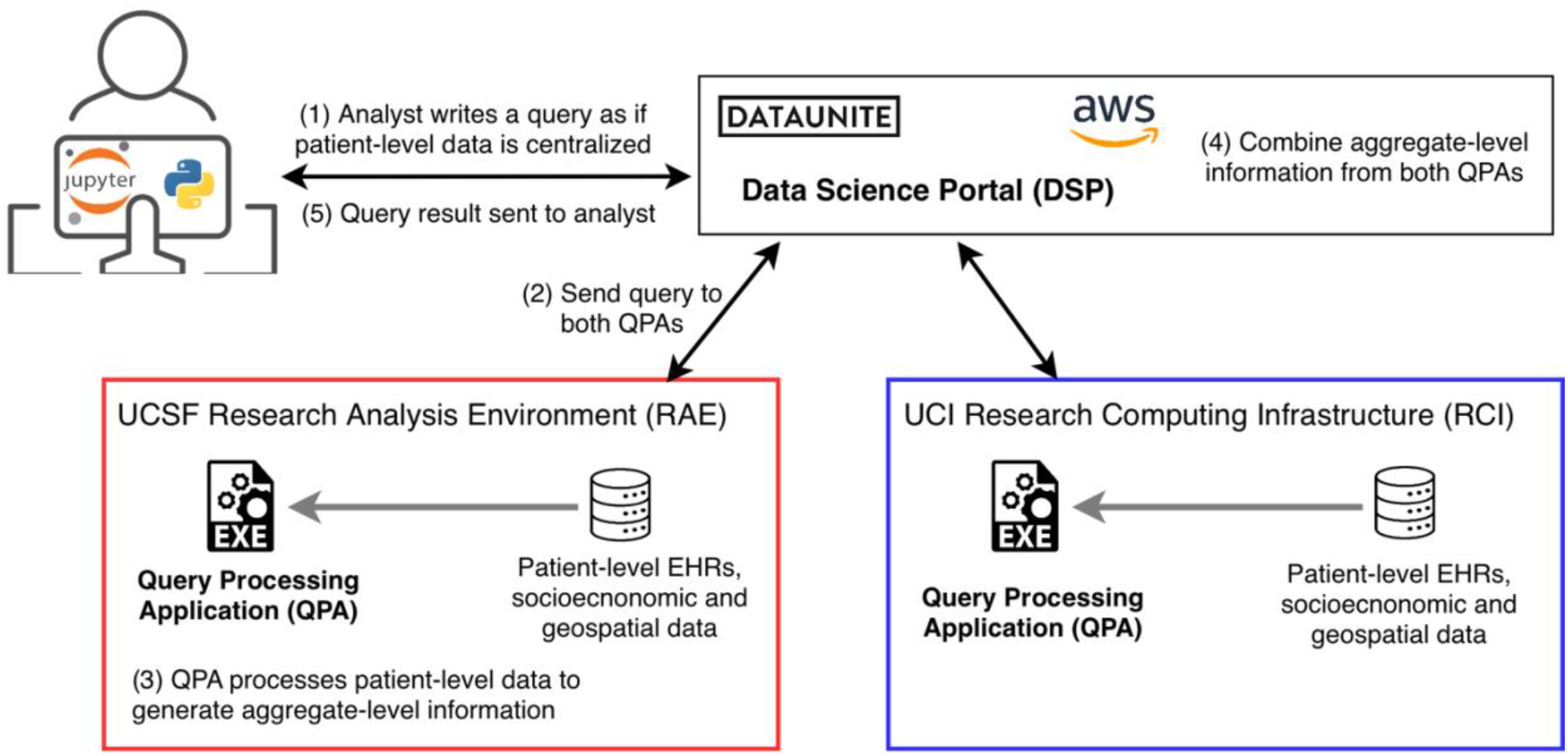
Virtual Pooling (VP) workflow and deployment at UCSF and UCI.

#### 1. Data Science Portal (DSP)

The DSP serves as the single/primary user interface through which study investigators and research personnel write Python code to perform data cleaning, preprocessing, harmonization, statistical analysis, and machine learning tasks. Analysts iteratively develop and refine analyses by submitting queries, reviewing results, and adjusting subsequent code as needed. We deployed the DSP on a public cloud (Amazon Web Services). Since the DSP operates outside secure health system environments, it receives and displays only aggregate-level results from the analyses, strictly prohibiting access to any patient-level data to maintain patient privacy.

#### 2. Query Processing Application (QPA)

We deployed the QPA, a lightweight software component, within each center’s secure environment—UCSF’s Research Analytics Environment (RAE) and UCI’s Research Computing Infrastructure (RCI). Each QPA receives analysis requests from the DSP, executes computations on the local patient-level data, and returns de-identified summary statistics or model weight updates. No patient-level data ever leaves a participating institution. This software component is compatible with commodity servers, both on-premises and cloud infrastructures, and does not require any changes to existing computing or networking configurations.

For this study, we implemented all analyses in Python using the VP platform v1.1, after the study-specific dataset was extracted from UCSF clinical data warehouse and supplied to the QPA at UCSF, and UCI’s study-specific dataset was extracted and provided to the UCI QPA. We additionally measured analyst-perceived latency, defined as the time between executing an analysis code segment and viewing its result. Below, we provide further details on the VP-specific workflow.

### Data Pre-Processing

We re-ran all data pre-processing, cleaning, and harmonization steps conducted in the original study, via the VP framework. The main operations we performed included:

#### Cleaning

The cleaning included renaming columns to a standardized format, converting to a homogeneous data type, and joining different data tables. Categorical variables, including race, ethnicity, and insurance payer type, were harmonized using user-defined recoding schemes to match across centers.

#### Feature Engineering

We computed derived features/covariates as in the original study through conditional logic and engineering target variables such as 1-year referral and 1-year visit completion based on structured logic applied to referral, eye visit, and primary care visit dates across centers.

#### Missing Values Imputation

We used multiple imputation via chained equations (MICE) to address missing data. Unlike conventional approaches that perform imputation independently at each center, VP allows analysts to choose between site-specific imputation or imputation based on a unified view of the multicenter data. Implementing MICE with a global data view in a federated setting is technically challenging, as MICE typically requires iterative access to the full dataset. The VP framework addresses this challenge by decomposing the imputation procedure into local center-level computations on cleaned data partitions, followed by secure aggregation coordinated through the DSP. We used this global imputation procedure in VP as it produces imputations consistent with those over a centralized dataset.

#### One-Hot Encoding

One-hot encoding was applied to categorical variables using a unified encoding scheme derived from the union of all observed category levels across participating centers. Thus, each center used an identical feature space, even when certain categories were present at only one center or were unevenly distributed across centers. By enforcing consistent feature representations across both centers, this approach enabled downstream modeling and inference to be performed seamlessly using the VP framework, without introducing site-specific feature mismatches.

### Statistical Analyses

We conducted three categories of statistical analysis. (A) Descriptive statistics summarized patient characteristics across sites. Categorical variables were reported as frequencies and percentages; continuous variables as means (standard deviations) or medians (interquartile ranges). Differences between sites were assessed using standardized mean differences (SMDs). (B) Univariate logistic regression was used to evaluate the association between each covariate and completion of a diabetic eye screening within one year. (C) Propensity score matching was used to estimate the average treatment effect (ATE) of receiving an automated referral on completion of diabetic eye screening. Although the original study used targeted maximum likelihood estimation (TMLE), VP v1.1 did not support TMLE but did support propensity score– based causal inference, and we therefore adopted this approach. Propensity scores were estimated via logistic regression with referral status as the treatment variable, and the ATE was computed as the mean difference in screening completion rates between matched groups. All p-values were two-sided, with statistical significance defined as *p* < 0.05.

## Results

Table 1 summarizes the key results of this study, including concordance between VP and the original centralized analysis across cohort construction, data preprocessing, statistical modeling, and causal inference, as well as analyst-perceived execution latency. We first report results related to VP’s deployability, followed by detailed validation of analytical concordance.

**Table 1.** Comparison of outputs from VP and centralized pooled analysis (Ayati et al.^27^), demonstrating exact reproduction of pooled estimates, preservation of site-specific heterogeneity, and interactive performance across the end-to-end workflow. Agreement is expressed as the number of identical outputs out of the total evaluated. Identical refers to numerical equality up to the 6^th^ decimal place.

|  |  |  |  |
| --- | --- | --- | --- |
| Validation Domain | Analysis Output | Result | Latency |
| **Data preprocessing and cohort construction** | Final analytical study cohort | Identical cohort (N = 8,240; UCSF = 2,592; UCI = 5,648) | <1s per step |
| **Exact reproduction of pooled estimates** | Baseline covariate descriptive statistics | 30/30 identical | <1s |
|  | Logistic regression odds ratios | 20/20 identical | <10s |
|  | Logistic regression *p*-values | 20/20 identical | <10s |
|  | Propensity score-based average treatment effect (ATE) of automated referrals | Identical point estimates | <30s |
| **Preservation of center-specific heterogeneity** | Standardized mean differences (UCSF & UCI) | 30/30 identical | <1s |

### Deployment and workflow feasibility

We deployed the QPA component of VP within the UCSF and UCI computing environments without requiring infrastructure changes, new or non-standard governance agreements, or dedicated IT support. Institutional security approvals were completed within 30 days at UCSF and 32 days at UCI, representing a relatively rapid deployment timeline for multicenter studies, and the QPAs were deployed on commodity computing resources. The entire study was executed end-to-end through this deployment, with the data analyst interacting exclusively with the DSP’s single analytical interface, abstracted away from—and effectively unaware of—the distributed multicenter system and its coordination mechanisms underneath.

### Execution latency

The VP framework introduced latency between query initiation and result display due to network communication required to transfer queries and results between the QPA instances deployed at the health systems and the coordinating DSP. For preprocessing and descriptive statistics, latency per operation was under one second, preserving an interactive experience. Latency increased for logistic regression because the algorithm required multiple rounds of communication between the QPA and DSP, though this complexity was fully abstracted/hidden from the analyst. Even so, logistic regression completed in under 10 seconds, with runtime dependent on the number of iterations needed for convergence. Propensity score matching involved additional internal steps and completed in under 30 seconds.

### Data pre-processing concordance

VP supported all preprocessing steps required for the study, including data cleaning, harmonization, missing value imputation, and one-hot encoding of categorical variables. These operations were executed through the DSP’s single, interactive analytical interface, allowing the analyst to iteratively inspect intermediate results and refine decisions without manual coordination with participating centers. Despite the heterogeneity of the underlying EHR data, VP produced consistent and harmonized feature representations across the two centers, enabling downstream analyses to proceed seamlessly. After using the same inclusion and exclusion criteria as the original study, the final study cohort consisted of 8,240 patients (2,592 from UCSF; 5,648 from UCI), the same as the original study^27^.

### Concordance of Statistical Estimation and Hypothesis Testing

As summarized in Table 1, descriptive statistics for all 30 covariates were numerically identical between the original and VP analyses, including means, standard deviations, medians, interquartile ranges, and category proportions (Supplementary Table 1 shows detailed results). Across both centers, 43% of patients received a referral for diabetic eye screening, but only 16% completed a screening visit within one year. Screening completion at UCSF and UCI was 21% and 13%, respectively. The SMDs calculated through VP were also identical to those from the original study^27^. As in the original study, SMDs exceeded 0.5 for over half of the key variables (neighborhood median income, insurance coverage type, and race/ethnicity), confirming substantial demographic and clinical heterogeneity between the two participating centers.

Univariate logistic regression (VP vs. original study) also yielded identical results. All 20 effect estimates, confidence intervals, and significance levels matched the original analysis (Supplementary Tables 2 and 3 show detailed results). The strongest univariate predictor of screening completion was receiving a referral (OR = 56.7; 95% CI: 42.1-76.4), followed by a history of other eye disease (OR = 6.4; 95% CI: 5.6-7.4). Higher HbA1c and BMI were associated with lower odds of screening, while older age and higher Charlson Comorbidity Index increased screening likelihood.

Finally, we estimated the ATE of automated referrals on screening completion using a propensity-matched cohort defined using VP. Center-specific and pooled estimates generated using VP were identical to those obtained from centrally run propensity score analyses on the pooled dataset from the original study. At UCSF, automated referrals were associated with an increase in screening completion from 21% to 36%; at UCI, completion increased from 13% to 34%; and, when data from both sites were analyzed jointly using VP, screening completion increased from 16% to 34%. These causal inference results are summarized in Table 1 and reported in detail in Supplementary Table 4.

These results were strongly consistent with the original paper which used TMLE for causal inference. TMLE is not currently supported by VP due to its underlying complexity. Nonetheless, these results matched the direction and magnitude of the original study, which reported increases from 21% to 34% at UCSF, 13% to 22% at UCI, and 16% to 28% overall. Across analyses (VP vs. original, and propensity score-based vs. TMLE), automated referrals consistently increased screening completion at both sites and in the pooled cohort, with comparable effect sizes and similar center-specific patterns. Together, these findings demonstrate that VP can recover both global treatment effects and center-specific heterogeneity without requiring patient-level data transfer.

## Discussion

Advances in artificial intelligence and the increasing use of multimodal healthcare data have intensified the need for large, diverse, and geographically distributed datasets, while simultaneously elevating concerns around data privacy. In parallel, countries including Canada, Singapore, India, and China have recently strengthened data protection regulations for personal health information^28–30^. Together, these trends point to a growing role for privacy-enhancing technologies (PETs) as a foundation for the next generation of scalable, collaborative, and privacy-preserving healthcare AI.

This study deployed and validated Virtual Pooling (VP), a new multicenter study execution platform, across two academic health systems—UCSF and UCI—and demonstrated that the platform is practical and can accurately replicate a multicenter retrospective study end-to-end with all the typical steps such as data cleaning, harmonization, descriptive statistics, regression, and causal inference—without requiring transfer of patient-level data from one center to the other. All results generated via VP exactly matched those from the original study, confirming the analytic fidelity of VP across diverse patient populations and EHR systems. This study also demonstrated that VP is easy to use, introduces modest execution latency, and requires no infrastructure changes or dedicated IT support.

Our findings have significant implications for the future of multi-institutional clinical research. Traditional data pooling remains the gold standard for complex statistical modeling, but is often infeasible due to privacy, legal, and operational constraints. VP overcomes these challenges by bringing the analysis to the data—allowing sites to retain complete control over sensitive patient information while still participating in high-impact, multicenter studies.

Importantly, this study involved moderately complex analysis: from data preprocessing and harmonization to multivariate regression and propensity score-based causal inference. Such pipelines are not natively supported by existing federated learning systems. VP brings extensive functionality while requiring no compromise in data security, governance alignment, or infrastructure requirements.

Furthermore, this study reaffirms the dual utility of federated analytics: preserving patient privacy while enabling rigorous causal discovery and policy-relevant insights. By aligning with results generated from a centralized dataset, the VP platform offers a more secure alternative for identifying care gaps^31–34^ and informing local intervention strategies to improve patient outcomes, rather than transferring patient-level data across institutions.

### Strengths

1. This study demonstrates that VP fills a critical gap in existing federated analytics approaches by enabling end-to-end execution of multicenter EHR studies starting from uncleaned and unharmonized data. Using VP, complex multi-step statistical workflows—including data cleaning, feature engineering, multiple imputation, one-hot encoding, univariate regression, and propensity score matching—can be executed in a federated environment while preserving patient-level data behind each center’s firewalls. Together, these results show that rigorous, real-world clinical analyses can be conducted without centralizing patient-level data.
2. The study leverages a previously published analysis over a pooled dataset as a benchmark, enabling direct comparison between pooled and federated approaches. This head-to-head validation provides strong external credibility, confirming that VP achieves equivalence to traditional centralized methods, including accurate effect estimation and detection of site-level heterogeneity. As a result, VP offers a more secure alternative to creating centralized datasets.
3. Finally, the study demonstrates that VP is practical to deploy and use in real-world settings: it is compatible with commodity infrastructure available at academic health systems, does not require infrastructure changes, non-standard governance agreements, or dedicated IT support, and enables analysts to conduct interactive, iterative analyses through a single interface without exposure to deployment or coordination complexity.

### Limitations

While this study successfully demonstrates the practicality of VP, it is limited to a demonstration using structured EHR data only. At the same time, a significant portion of clinical knowledge is captured in unstructured data sources such as clinical notes. Incorporating free-text notes or other unstructured data sources, such as images or genomic data, and demonstrating VP’s practicality for these data domains is future work. In addition, VP currently relies on propensity score–based methods for causal effect estimation; incorporating additional causal inference algorithms and estimation frameworks such as TMLE to broaden its analytical capabilities is planned for future versions. Furthermore, while this study validated VP for a two-center study, future work would evaluate VP’s scalability for studies involving a larger consortium with more centers.

### Conclusion

This work provides strong empirical validation that VP enables practical and accurate federated analysis of EHR data while supporting the full end-to-end lifecycle of a clinical study, from data cleaning and harmonization to advanced statistical analysis. VP’s single analytical interface allows researchers to execute complex, multi-step workflows without manual site coordination or specialized infrastructure expertise. By lowering both technical and operational barriers, this study lays the foundation for a new class of multicenter clinical research that is not only privacy-preserving but also easy to execute in real-world settings where data sharing is time-consuming, paperwork-intensive, or restricted. Broad adoption of VP could transform the landscape of real-world evidence generation across diverse healthcare institutions by facilitating the inclusion of more diverse and heterogeneous datasets, accelerating discovery across population health and precision medicine while preserving institutional autonomy and patient trust.

## Data Availability

This study used deidentified electronic medical record (EMR) data from the University of California, San Francisco (UCSF) and the University of California, Irvine (UCI). The study protocol was approved by the UCSF Institutional Review Board (IRB #23-40187) and determined to be non-human subjects research by the UCI IRB. Due to institutional and patient privacy restrictions, the data are not publicly available but may be obtained from the corresponding author upon reasonable request and completion of a data use agreement with UCSF and UCI.

