## Supplementary Tables for "Virtual Pooling Enables Accurate, End-to-End Multi-Institutional Study Execution and Causal Inference Without Centralized Data Sharing"

### Supplementary Information

This document presents comparisons of all covariates and outcomes between the original study by Ayati et al.[27] and the analysis done through VP in this study. Supplementary Table 1 reports descriptive statistics and standardized mean differences (SMDs) for all 30 variables from the original study; Supplementary Tables 2 and 3 summarize logistic regression odds ratios and corresponding p-values for the subset of variables examined in the original study's regression analysis; and Supplementary Table 4 summarizes average treatment effect sizes of automated referrals on screening completion.

**Supplementary Table 1: Baseline cohort characteristics from Ayati et al.[27] versus this study via VP. Results are presented for UCSF+UCI combined cohort and stratified by center.**

| | Combined (n = 8240) | Combined VP (n=8240) | UCSF (n = 2592) | UCSF VP (n = 2592) | UCI (n = 5648) | UCI VP (n = 5648) | SMD | SMD VP |
|---|---|---|---|---|---|---|---|---|
| **Demographics** | | | | | | | | |
| Age at Visit (Mean, SD) | 54.9 (12.2) | 54.9 (12.2) | 57.1 (11.9) | 57.1 (11.9) | 53.8 (12.1) | 53.8 (12.1) | 0.275 | 0.275 |
| **Sex (n, %)** | | | | | | | | |
| Female | 4246 (51.5) | 4246 (51.5) | 1277 (49.3) | 1277 (49.3) | 2969 (52.6) | 2969 (52.6) | 0.066 | 0.066 |
| Male | 3994 (48.5) | 3994 (48.5) | 1315 (50.7) | 1315 (50.7) | 2679 (47.4) | 2679 (47.4) | | |
| **Race/Ethnicity (n, %)** | | | | | | | | |
| Asian | 1716 (20.8) | 1716 (20.8) | 922 (35.6) | 922 (35.6) | 794 (14.1) | 794 (14.1) | 1.102 | 1.102 |
| Black or African American | 483 (5.9) | 483 (5.9) | 342 (13.2) | 342 (13.2) | 141 (2.5) | 141 (2.5) | | |
| Hispanic or Latino | 3734 (45.3) | 3734 (45.3) | 380 (14.7) | 380 (14.7) | 3354 (59.4) | 3354 (59.4) | | |
| Indigenous | 121 (1.5) | 121 (1.5) | 63 (2.4) | 63 (2.4) | 58 (1.0) | 58 (1.0) | | |
| White | 1640 (19.9) | 1640 (19.9) | 635 (24.5) | 635 (24.5) | 1005 (17.8) | 1005 (17.8) | | |
| **Insurance Type (n, %)** | | | | | | | | |
| Commercial | 2623 (31.8) | 2623 (31.8) | 1041 (40.2) | 1041 (40.2) | 1582 (28.0) | 1582 (28.0) | 1.012 | 1.012 |
| Medicaid | 2392 (29.0) | 2392 (29.0) | 436 (16.8) | 436 (16.8) | 1956 (34.6) | 1956 (34.6) | | |
| Medicare | 1723 (20.9) | 1723 (20.9) | 813 (31.4) | 813 (31.4) | 910 (16.1) | 910 (16.1) | | |
| Self-Pay | 1207 (14.6) | 1207 (14.6) | 27 (1.0) | 27 (1.0) | 1180 (20.9) | 1180 (20.9) | | |
| **Clinical Characteristics** | | | | | | | | |
| A1C Level (Median, [IQR]) | 7.4 [6.5,8.7] | 7.4 [6.5,8.7] | 7.1 [6.4,8.1] | 7.1 [6.4,8.1] | 7.5 [6.6,9.1] | 7.5 [6.6,9.1] | -0.17 | -0.17 |
| BMI (Mean, SD) | 31.4 (6.5) | 31.3 (6.5) | 30.8 (6.6) | 30.8 (6.6) | 31.5 (6.4) | 31.5 (6.4) | -0.111 | -0.111 |

| | | | | | | | | |
|---|---|---|---|---|---|---|---|---|
| Charlson Score (Median, [IQR]) | 2.0 [1.0,4.0] | 2.0 [1.0,4.0] | 1.0 [1.0,3.0] | 1.0 [1.0,3.0] | 2.0 [1.0,4.0] | 2.0 [1.0,4.0] | -0.144 | -0.144 |
| History of eye diseases (n, %) | 3476 (42.2) | 3476 (42.2) | 885 (34.1) | 885 (34.1) | 2591 (45.9) | 2591 (45.9) | -0.241 | -0.241 |
| Insulin Use (n, %) | 1857 (22.5) | 1857 (22.5) | 458 (17.7) | 458 (17.7) | 1399 (24.8) | 1399 (24.8) | -0.174 | -0.174 |
| **PCP Specialty (n, %)** | | | | | | | | |
| Family Medicine | 4082 (49.5) | 4082 (49.5) | 376 (14.5) | 376 (14.5) | 3706 (65.6) | 3706 (65.6) | 1.264 | 1.264 |
| Internal Medicine | 1973 (23.9) | 1973 (23.9) | 1097 (42.3) | 1097 (42.3) | 876 (15.5) | 876 (15.5) | | |
| Primary Care | 2056 (25.0) | 2056 (25.0) | 1105 (42.6) | 1105 (42.6) | 951 (16.8) | 951 (16.8) | | |
| **Location Based Variables (Median, [IQR]):** | | | | | | | | |
| ADI Status Rank | 5.0 [2.0,6.0] | 5.0 [2.0,6.0] | 2.0 [1.0,2.0] | 2.0 [1.0,2.0] | 5.0 [4.0,6.0] | 5.0 [4.0,6.0] | -0.989 | -0.989 |
| Driving Distance | 13.6 [7.5,20.3] | 13.6 [7.5,20.3] | 4.6 [3.4,14.1] | 4.6 [3.4,14.1] | 16.9 [11.2,20.9] | 16.9 [11.2,20.9] | 0.069 | 0.069 |
| Median Income | 95.6 [81.6,128.3] | 95.0 [81.0,128.0] | 126.9 [104.5,152.1] | 126.9 [104.5,152.1] | 89.7 [80.3,114.4] | 89.0 [80.0,114.0] | 1.033 | 1.033 |
| Poverty Rate | 10.3 [7.7,14.8] | 10.3 [7.7,14.8] | 9.0 [6.6,13.1] | 9.0 [6.6,13.1] | 10.7 [8.8,15.1] | 10.7 [8.8,15.1] | -0.293 | -0.293 |
| Gini Index | 0.4 [0.4,0.5] | 0.4 [0.4,0.5] | 0.5 [0.4,0.5] | 0.5 [0.4,0.5] | 0.4 [0.4,0.4] | 0.4 [0.4,0.4] | 1.198 | 1.198 |
| No Internet Access Rate | 7.7 [5.6,9.8] | 7.7 [5.6,9.8] | 7.0 [6.1,8.5] | 7.0 [6.1,8.5] | 7.8 [5.6,10.0] | 7.8 [5.6,10.0] | -0.188 | -0.188 |
| Unemployment Rate | 3.6 [3.2,4.0] | 3.6 [3.2,4.0] | 3.6 [3.0,4.4] | 3.6 [3.0,4.4] | 3.6 [3.3,4.0] | 3.6 [3.3,4.0] | 0.077 | 0.077 |
| Vehicle Access | 5.5 [3.6,9.6] | 5.5 [3.6,9.6] | 15.0 [6.5,25.2] | 15.0 [6.5,25.2] | 4.6 [3.3,7.0] | 4.6 [3.3,7.0] | 1.128 | 1.128 |
| Single-Adult Household Rate | 19.3 [16.4,24.3] | 19.3 [16.4,24.3] | 26.9 [20.1,35.3] | 26.9 [20.1,35.3] | 17.9 [14.4,20.2] | 17.9 [14.4,20.2] | 1.232 | 1.232 |
| College Education Rate | 64.8 [48.3,78.9] | 64.8 [48.3,78.9] | 75.8 [65.6,85.1] | 75.8 [65.6,85.1] | 54.4 [44.8,71.2] | 54.4 [44.8,71.2] | 1.058 | 1.058 |

**Supplementary Table 2: Univariate logistic regression analysis for 1 year eye screening visits at UCSF and UCI.**

| | UCSF OR [95% CI] | UCSF VP OR [95% CI] | UCSF p-value | UCSF VP p-value | UCI OR [95% CI] | UCI VP OR [95% CI] | UCI p-value | UCI VP p-value |
|---|---|---|---|---|---|---|---|---|
| **Demographics** | | | | | | | | |
| Age at Visit | 1.0 [1.0, 1.0] | 1.0 [1.0, 1.0] | 0.013 | 0.013 | 1.0 [1.0, 1.1] | 1.0 [1.0, 1.1] | <0.001 | <0.001 |
| Female | 1.2 [1.0, 1.5] | 1.2 [1.0, 1.5] | 0.023 | 0.023 | 0.9 [0.8, 1.0] | 0.9 [0.8, 1.0] | 0.094 | 0.094 |
| **Race/Ethnicity** | | | | | | | | |
| Asian | 1.2 [1.0, 1.5] | 1.2 [1.0, 1.5] | 0.076 | 0.076 | 2.7 [2.3, 3.3] | 2.7 [2.3, 3.3] | <0.001 | <0.001 |
| Black or African American | 1.2 [0.9, 1.6] | 1.2 [0.9, 1.6] | 0.17 | 0.17 | 1.2 [0.8, 1.9] | 1.2 [0.8, 1.9] | 0.434 | 0.434 |
| Hispanic or Latino | 1.0 [0.8, 1.3] | 1.0 [0.8, 1.3] | 0.966 | 0.966 | 0.3 [0.3, 0.4] | 0.3 [0.3, 0.4] | <0.001 | <0.001 |
| Indigenous | 1.4 [0.8, 2.5] | 1.4 [0.8, 2.5] | 0.229 | 0.229 | 1.7 [0.9, 3.2] | 1.7 [0.9, 3.2] | 0.104 | 0.104 |

| | | | | | | | | |
|---|---|---|---|---|---|---|---|---|
| White | 0.6 [0.5, 0.7] | 0.6 [0.5, 0.7] | <0.001 | <0.001 | 1.8 [1.5, 2.2] | 1.8 [1.5, 2.2] | <0.001 | <0.001 |
| **Insurance Status** | | | | | | | | |
| Commercial | 0.4 [0.4, 0.5] | 0.4 [0.4, 0.5] | <0.001 | <0.001 | 3.3 [2.8, 3.8] | 3.3 [2.8, 3.8] | <0.001 | <0.001 |
| Medicaid | 2.8 [2.2, 3.5] | 2.8 [2.2, 3.5] | <0.001 | <0.001 | 0.1 [0.1, 0.2] | 0.1 [0.1, 0.2] | <0.001 | <0.001 |
| Medicare | 1.0 [0.8, 1.2] | 1.0 [0.8, 1.2] | 0.891 | 0.891 | 4.6 [3.8, 5.4] | 4.6 [3.8, 5.4] | <0.001 | <0.001 |
| Self-Pay | 0.9 [0.3, 2.3] | 0.9 [0.3, 2.3] | 0.762 | 0.762 | 0.1 [0.0, 0.1] | 0.1 [0.0, 0.1] | <0.001 | <0.001 |
| **Clinical Characteristics** | | | | | | | | |
| A1C Level | 1.0 [0.9, 1.1] | 1.0 [0.9, 1.1] | 0.874 | 0.874 | 0.9 [0.9, 1.0] | 0.9 [0.9, 1.0] | <0.001 | <0.001 |
| BMI | 1.0 [1.0, 1.0] | 1.0 [1.0, 1.0] | 0.197 | 0.197 | 1.0 [1.0, 1.0] | 1.0 [1.0, 1.0] | 0.003 | 0.003 |
| Charlson Score | 1.1 [1.0, 1.1] | 1.1 [1.0, 1.1] | 0.021 | 0.021 | 1.2 [1.2, 1.2] | 1.2 [1.2, 1.2] | <0.001 | <0.001 |
| Eye Diseases | 2.7 [2.2, 3.3] | 2.7 [2.2, 3.3] | <0.001 | <0.001 | 40.7 [28.0, 59.3] | 40.7 [28.0, 59.3] | <0.001 | <0.001 |
| Screening Referral in 1 Year | 21.9 [15.1, 31.9] | 21.9 [15.1, 31.9] | <0.001 | <0.001 | 126.7 [75.7, 212.1] | 126.7 [75.7, 212.1] | <0.001 | <0.001 |
| Insulin | 1.2 [0.9, 1.5] | 1.2 [0.9, 1.5] | 0.149 | 0.149 | 0.6 [0.5, 0.7] | 0.6 [0.5, 0.7] | <0.001 | <0.001 |
| **Provider Specialty** | | | | | | | | |
| Family Medicine | 1.2 [0.9, 1.5] | 1.2 [0.9, 1.5] | 0.243 | 0.243 | 0.2 [0.2, 0.2] | 0.2 [0.2, 0.2] | <0.001 | <0.001 |
| Internal Medicine | 1.1 [0.9, 1.4] | 1.1 [0.9, 1.4] | 0.17 | 0.17 | 5.2 [4.4, 6.1] | 5.2 [4.4, 6.1] | <0.001 | <0.001 |
| Primary Care | 0.8 [0.7, 1.0] | 0.8 [0.7, 1.0] | 0.044 | 0.044 | 2.2 [1.9, 2.7] | 2.2 [1.9, 2.7] | <0.001 | <0.001 |

**Supplementary Table 3: Univariate logistic regression analysis for 1 year eye screening visits for both sites combined (UCSF+UCI).**

| | Combined OR [95% CI] | Combined VP OR [95% CI] | Combined p-value | Combined VP p-value |
|---|---|---|---|---|
| **Demographics** | | | | |
| Age at Visit | 1.0 [1.0, 1.0] | 1.0 [1.0, 1.0] | <0.001 | <0.001 |
| Female | 1.0 [0.9, 1.1] | 1.0 [0.9, 1.1] | 0.888 | 0.888 |
| **Race/Ethnicity** | | | | |
| Asian | 2.0 [1.8, 2.3] | 2.0 [1.8, 2.3] | <0.001 | <0.001 |
| Black or African American | 1.5 [1.2, 1.9] | 1.5 [1.2, 1.9] | 0.001 | 0.001 |
| Hispanic or Latino | 0.4 [0.3, 0.4] | 0.4 [0.3, 0.4] | <0.001 | <0.001 |
| Indigenous | 1.7 [1.1, 2.6] | 1.7 [1.1, 2.6] | 0.013 | 0.013 |
| White | 1.2 [1.1, 1.4] | 1.2 [1.1, 1.4] | 0.008 | 0.008 |
| **Insurance Status** | | | | |
| Commercial | 1.6 [1.4, 1.8] | 1.6 [1.4, 1.8] | <0.001 | <0.001 |
| Medicaid | 0.5 [0.4, 0.5] | 0.5 [0.4, 0.5] | <0.001 | <0.001 |
| Medicare | 2.6 [2.3, 2.9] | 2.6 [2.3, 2.9] | <0.001 | <0.001 |
| Self-Pay | 0.1 [0.0, 0.1] | 0.1 [0.0, 0.1] | <0.001 | <0.001 |
| **Clinical Characteristics** | | | | |
| A1C Level | 0.9 [0.9, 1.0] | 0.9 [0.9, 1.0] | <0.001 | <0.001 |
| BMI | 1.0 [1.0, 1.0] | 1.0 [1.0, 1.0] | <0.001 | <0.001 |
| Charlson Score | 1.1 [1.1, 1.2] | 1.1 [1.1, 1.2] | <0.001 | <0.001 |
| Eye Diseases | 6.5 [5.6, 7.4] | 6.5 [5.6, 7.4] | <0.001 | <0.001 |
| Screening Referral in 1 Year | 56.7 [42.1, 76.4] | 56.7 [42.1, 76.4] | <0.001 | <0.001 |
| Insulin | 0.7 [0.6, 0.9] | 0.7 [0.6, 0.9] | <0.001 | <0.001 |
| **Provider Specialty** | | | | |
| Family Medicine | 0.3 [0.2, 0.3] | 0.3 [0.2, 0.3] | <0.001 | <0.001 |
| Internal Medicine | 2.8 [2.5, 3.2] | 2.8 [2.5, 3.2] | <0.001 | <0.001 |
| Primary Care | 1.6 [1.4, 1.8] | 1.6 [1.4, 1.8] | <0.001 | <0.001 |

**Supplementary Table 4: Screening completion rates with and without automated referrals from Ayati et al.[27] versus VP.**

| Cohort | VP analysis (PS-based ATE) | Original study (PS-based ATE) | Original study (TMLE) |
|---|---|---|---|
| UCSF | 21% → 36% | 21% → 36% | 21% → 34% |
| UCI | 13% → 34% | 13% → 34% | 13% → 22% |
| Combined | 16% → 34% | 16% → 34% | 16% → 28% |
